# Objective sleep parameters and diurnal blood pressure in concurrent hypertension and type 2 diabetes

**DOI:** 10.1101/2025.10.20.25338423

**Authors:** Yan Zhao, Yuzhe Zha, Yuchan Zheng, Lijun Wei, Baoyi Chen, Zefan Zhang, Xiao Tan

## Abstract

**Background:** Hypertension is a primary cardiovascular complication in type 2 diabetes, with a significantly increased risk of morbidity and mortality compared to the general population. Ambulatory blood pressure monitoring is essential because blood pressure exhibits circadian rhythmicity, and sleep plays a crucial role in modulating nocturnal blood pressure patterns and overall cardiovascular risk.

**Method:** A total of 20 patients (63.75 ± 4.44 years old, 40% female, duration of T2D: 12.3 ± 6.24 years) underwent ambulatory blood pressure monitoring (ABPM) and actigraphy in a free-living condition. Multi-day sleep parameters including total sleep time, sleep efficiency, wake after sleep onset, sleep onset latency and number of awakenings were assessed by wrist actigraphy supplemented with sleep diary, ABPM was measured for 24 hours during the sleep assessment period. Associations between parameters and ABPM across total sleep measurement period and the exact date of ABPM measurement were investigated, respectively.

**Results:** Sleep onset latency was associated with 24-hour coefficient of variance of systolic blood pressure (β-coefficient: 0.323, 95% CI: [0.014, 0.632], p = 0.04). Inter-day sleep efficiency was inversely associated with SD and CV (SD: β-coefficient: -0.373, 95% CI: [-0.676, -0.071], P = 0.02; CV: β-coefficient: -0.588, 95% CI: [-1.141, -0.036], P = 0.04) of diastolic blood pressure.

**Conclusion:** In individuals with T2D and hypertension, both acute and chronic sleep disturbances, particularly prolonged sleep onset latency and reduced multi-day sleep efficiency, are closely associated with increased blood pressure variability, highlighting the importance of sleep quality in cardiovascular risk management.

## 1. Introduction

Type 2 diabetes (T2D) presents substantial challenges to healthcare systems due to its wide-ranging complications. Among these complications, cardiovascular diseases are particularly concerning, with hypertension being a major contributor to adverse outcomes including atherosclerosis, vascular inflammation and dyslipidemia (1). Previous research has shown that genetically instrumented T2D significantly increases the risk of developing hypertension, underscoring the importance of managing blood pressure in T2D patients and the necessity of regular monitoring of blood pressure (2). Population-based studies have also reported similar findings, with approximately 50% to 80% of T2D patients in the U.S. suffering from hypertension (3). Furthermore, a study revealed that by the age of fifty, 85% of individuals with T2D are diagnosed with hypertension (4). This number is expected to rise in the coming decades as the population continues to age.

Blood pressure exhibits 24-hour circadian pattern, which is typically higher during the day and lower at night. Non-dipping blood pressure pattern, typically defined as night-day ambulatory blood pressure ratio ≥0.9 (5), is a risk factor for coronary artery disease, heart failure (6), left ventricular hypertrophy, and other cardiovascular complications (7, 8). Among patients with T2D, the non-dipping blood pressure pattern is more prevalent than the dipping pattern and has been associated with an elevated risk of cardiovascular events and increased mortality (9, 10). Sleep has been identified as an important factor related with diurnal BP pattern (11). On the other hand, T2D patients are prone to various sleep and circadian disorders including but not limited to insomnia, obstructive sleep apnea, and unregulated chronotype (12). Disrupted sleep patterns are associated with heightened sympathetic nervous system activity and diminished parasympathetic tone, leading to elevated nocturnal BP and a blunted dipping profile (13), and inadequate or fragmented sleep also contributes to endothelial dysfunction, characterized by reduced vasodilatory capacity and increased arterial stiffness, which further promotes hypertension (14). Moreover, irregular sleep can disrupt the circadian rhythm, impairing the temporal regulation of blood pressure across the day-night cycle (15). Understanding the interaction between sleep and diurnal blood pressure is of clinical relevance for improving the management of T2D and reducing the risk of cardiovascular complications. Of note, diabetes-related sleep disruptions may further exacerbate blood pressure dysregulation, creating a vicious cycle that deteriorates both sleep quality and cardiovascular function. Understanding the complex interplay between sleep and blood pressure control in T2D patients is therefore essential for developing targeted interventions to mitigate cardiovascular risk.

Ambulatory blood pressure monitoring (ABPM) is a technique that continuously records blood pressure, offering a more comprehensive assessment compared to clinic measurements. It provides detailed insights into diurnal blood pressure patterns and enables the diagnosis of conditions that may be missed in clinical settings, such as white-coat hypertension, masked hypertension, and nocturnal hypertension (16, 17). Identifying these hypertension subtypes is crucial for assessing cardiovascular risk more accurately. Meanwhile, actigraphy serves as a non-invasive and reliable method for objectively measuring sleep-wake cycles and activity levels over extended periods. By integrating ABPM with actigraphy, researchers can obtain a more holistic view of how blood pressure fluctuations interact with sleep. This combined approach enhances the understanding of sleep-related blood pressure dysregulation, which may inform more precise risk stratification and targeted interventions in this population.

Therefore, the present study aimed to investigate both the concurrent and inter-day associations between sleep and diurnal blood pressure in patients with T2D comorbid with hypertension. We hypothesized that disrupted objective sleep characterized by shorter sleep duration, lower sleep efficiency, and increased sleep fragmentation would be associated with elevated nocturnal blood pressure, a blunted nocturnal dipping pattern, and higher overall 24-hour blood pressure. Furthermore, we hypothesized that inter-day variability in sleep patterns would correlate with increased blood pressure variability and greater cardiovascular risk.

## 2. Methods

### 2.1 Study population

This was an observational study among adults with T2D comorbid with hypertension. Participants were involved through a clinical trial including T2D patients (ARRA, NCT06145542). Baseline measurement data from ARRA participants with self-reported hypertension (n=20) were incorporated into the analyses. Participants were recruited through the Maigaoqiao Community Health Service Center, Nanjing Municipality, China. T2D was diagnosed by physicians according to the following criteria: 1) random blood glucose ≥11.1 mmol/L, 2) fasting blood glucose ≥7.0 mmol/L, 3) 2-hour post-oral glucose tolerance test blood glucose ≥11.1 mmol/L, 4) HbA1c ≥ 48 mmol/mol (6.5%). Exclusion criteria were 1) type 1 diabetes, 2) present shift work, 3)severe kidney or liver disease, 4)diabetic ketoacidosis, 5)recurrent hypoglycemic events in the previous 3 months, 6)dementia and other mental disorders. Among the 65 participants in the ARRA trial, 20 participants were eligible for this study. The study protocol was approved by the ethics committee of the Affiliated Rehabilitation Hospital of Nanjing Sport Institute in accordance with the principles of the Declaration of Helsinki. Participation was voluntary, and a written consent form was obtained from each participant prior to the measurements.

### 2.2 Baseline characteristics

Baseline characteristics including age, sex, duration of diabetes, hypertension status were collected via printed questionnaire. Weight, height were measured at the community health center. BMI was calculated as weight (kg) divided by the square of height (m^2^). Physical activity is assessed via International Physical Activity Questionnaire (IPAQ), and IPAQ metabolic equivalent of task (MET) per week was calculated based on the standardized formula (18).

### 2.3 Measurement of objective sleep parameters

Participants were instructed to wear an actigraphy (GT3x, ActiGraph LLC, Pensacola, Fl, USA) on their non-dominant wrist for a consecutive of 14 days, and to keep wearing unless they were showering, bathing or swimming. A sampling frequency of 60Hz was set for determining acceleration. Participants’ bedtime and morning awakening time collected by sleep diary, as well as light intensity (in lux) determined by the actigraphy devices were used to assist actigraphy data scoring. The Cole-Kripke algorithm was applied to identify sleep and wake state in 1-minute epochs. Total sleep time (TST), sleep efficiency (SE), sleep onset latency (SL), wake after sleep onset (WASO), and number of awakenings of each night were obtained or calculated.

### 2.4 Ambulatory blood pressure monitoring

The ambulatory blood pressure monitoring (ABPM, Oscar 2 ABP monitor, Suntech Medical, Raleigh, NC, USA) was implemented under a 30-minute frequency for a 24-hour period (15:00 – 15:00) during the 14-day actigraphy measurement. The ABPM cuff was placed on the upper arm of the non-dominant side, and the monitor unit was attached to the waist on the opposite side.

Nighttime blood pressure (BP) was defined as ABPM readings recorded from onset to final awakening, based on the actigraphy data. All other readings outside this period were classified as daytime BP. 24-h systolic BP, daytime systolic BP, nighttime systolic BP, weighted SD of systolic BP, weighted CV of systolic BP, 24-h diastolic BP, daytime diastolic BP, nighttime diastolic BP, weighted SD of diastolic BP, weighted CV of diastolic BP and dipping ratio of each participant were obtained or calculated.

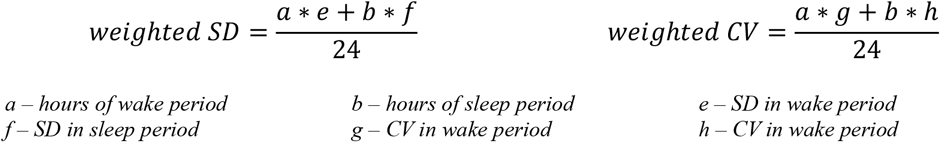

Nocturnal hypertension referred to average nighttime blood pressure ≥120/70 mm Hg. Nocturnal hypertension with wakeful blood pressure < 135/85 mm Hg was defined as isolated nocturnal hypertension and as uncontrolled nocturnal hypertension under antihypertensive medication (19, 20, 21). Additionally, nocturnal blood pressure patterns were classified into four categories based on the percentage of nighttime decline relative to daytime values. Normal dippers exhibited a 10-20% nocturnal reduction (5). Non-dippers showed less than 10% decline, while extreme dippers experienced more than 20% drop (5). In contrast, reverse dippers demonstrated higher nighttime than daytime blood pressure (5).

### 2.5 Statistical analyses

All continuous variables were expressed as mean ± standard deviation and categorical variables are expressed as frequencies (percentages). K-nearest neighbor imputation was used to address missing values. Linear regression was used to explore the relationship between sleep and blood pressure, adjusted for age, sex, BMI, the duration of diabetes, medication use, hypertension status and IPAQ MET. The results were shown as coefficient (95% CI), and statistical significance was indicated with asterisks. Analysis of covariance (ANCOVA) was conducted to investigate the relationship between hypertension type and objective sleep parameters. Statistical analyses were conducted using R (Version 4.4.1) and SPSS (Version 26.0, IBM Corp., Armonk, NY, USA). A two-tailed P value of less than 0.05 was considered as statistically significant.

## 3. Results

### 3.1 Sample descriptives

Descriptive statistics of the sample were displayed in **Table 1**. A total of 20 participants (8 female, 40%) participated in this study. Average age was 63.75 ± 4.44 years, mean BMI was 25.9 ± 2.63 kg/m^2^, and average duration of diabetes was 12.3 ± 6.2 years. Based on 14-days sleep monitoring, mean TST was 417.2 ± 56.0 minutes, mean WASO was 71.0 ± 23.4 minutes, and mean sleep efficiency was 84.1 ± 4.5%.

**Table 1.**
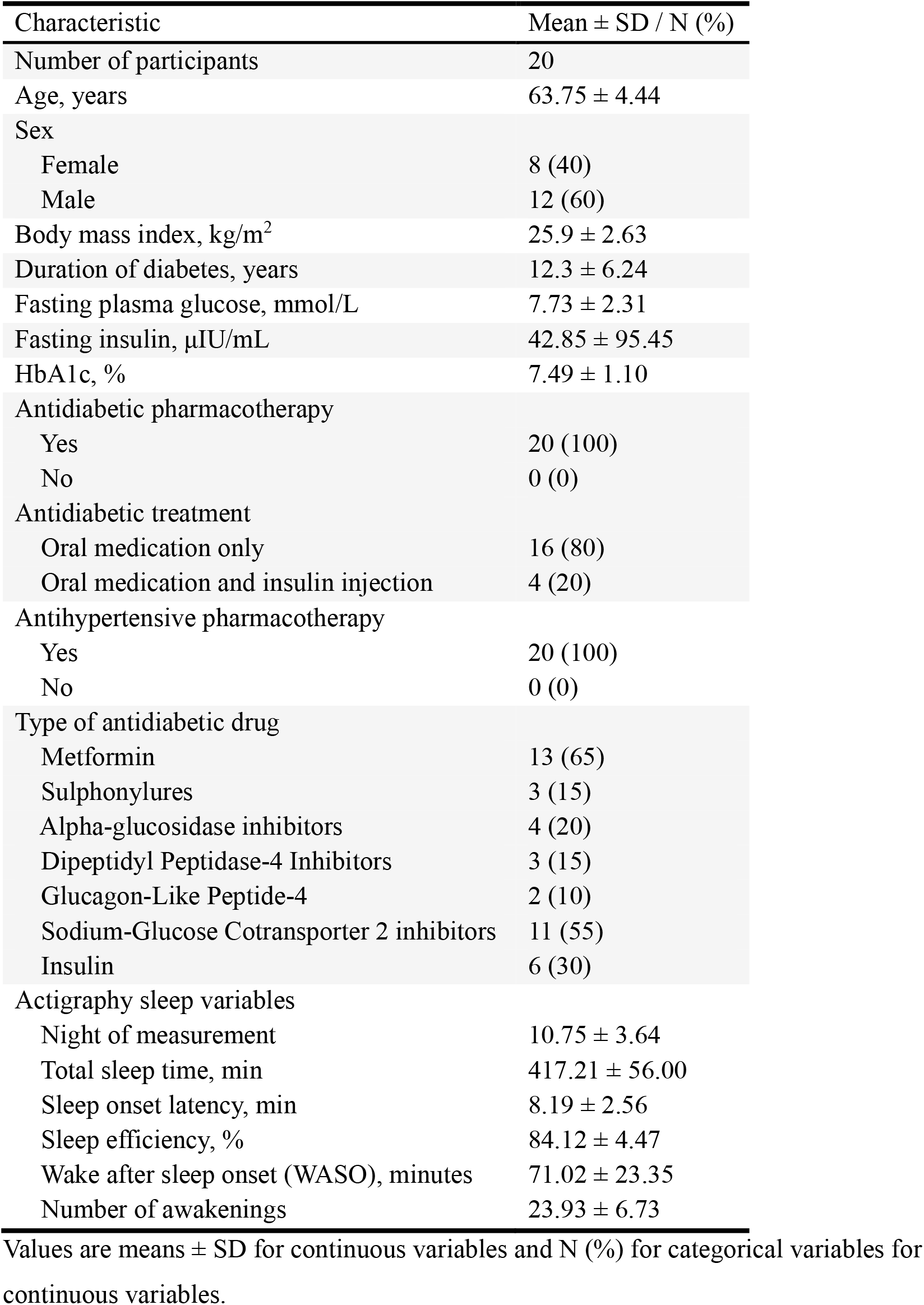
Sample Characteristics.

According to the ABPM results (**Table 2**), 40% of the participants had normal blood pressure, while 60% exhibited elevated blood pressure. 20% of the participants had a dipping ratio between 0.8 and 0.9, 60% had a ratio between 0.9 and 1, and 20% had a ratio greater than 1. Mean SBP of the ABPM period was 123.54 ± 11.33 mmHg (SD = 13.68 ± 3.55 mmHg, CV = 11.03 ± 2.48%), mean DBP was 75.29 ± 4.37 mmHg (SD = 10.47 ± 3.27 mmHg, CV = 13.93 ± 4.28%). Daytime SBP was 125.05 ± 11.49 mmHg (SD = 14.14 ± 4.26 mmHg, CV = 11.23 ± 2.93%), and daytime DBP was 77.22 ± 4.93 mmHg (SD = 10.38 ± 3.90 mmHg, CV = 13.46 ± 4.93%). Nighttime SBP was 118.64± 12.72 mmHg (SD = 9.67 ± 3.18 mmHg, CV = 8.27 ± 3.00%), and nighttime DBP was 69.14 ± 6.03 mmHg (SD = 6.51 ± 2.51 mmHg, CV = 9.70 ± 4.31%).

**Table 2.** ABPM variables.

| ABPM variables | Mean ± SD / N (%) |
| --- | --- |
| Arterial pressure | 91.04 ± 6.26 |
| Heart rate | 78.00 ± 8.00 |
| Hypertension types |  |
| Normal blood pressure | 8 (40) |
| Uncontrolled nocturnal hypertension | 8 (40) |
| Sustained hypertension | 4 (20) |
| Dipping pattern (SBP) |  |
| Normal dippers | 4 (20) |
| Non-dippers | 12 (60) |
| Reverse dippers | 4 (20) |
| SBP |  |
| Mean over 24 hours | 123.54 ± 11.33 |
| SD over 24 hours | 13.68 ± 3.55 |
| CV over 24 hours | 11.03 ± 2.48 |
| Mean in wake period | 125.05 ± 11.49 |
| SD in wake period | 14.14 ± 4.26 |
| CV in wake period | 11.23 ± 2.93 |
| Mean in sleep period | 118.64 ± 12.72 |
| SD in sleep period | 9.67 ± 3.18 |
| CV in sleep period | 8.27 ± 3.00 |
| Weighted SD | 12.70 ± 3.19 |
| Weighted CV | 10.26 ± 2.18 |
| DBP |  |
| Mean over 24 hours | 75.29 ± 4.37 |
| SD over 24 hours | 10.47 ± 3.27 |
| CV over 24 hours | 13.93 ± 4.28 |
| Mean in wake period | 77.22 ± 4.93 |
| SD in wake period | 10.38 ± 3.90 |
| CV in wake period | 13.46 ± 4.93 |
| Mean in sleep period | 69.14 ± 6.03 |
| SD in sleep period | 6.51 ± 2.51 |
| CV in sleep period | 9.70 ± 4.31 |
| Weighted SD | 9.17 ± 2.83 |
| Weighted CV | 12.24 ± 3.67 |

Values are means ± SD for continuous variables and N (%) for categorical variables for continuous variables.

### 3.2 Associations between concurrent sleep and ABPM parameters

The relationship between sleep characteristics and concurrently assessed ABPM parameters are demonstrated in **Table 3**. Sleep onset latency was correlated with 24-hour CV of SBP (β-coefficient: 0.323, 95% CI: [0.014, 0.632], p = 0.04). CV during the wake period of SBP was associated with number of awakenings (β-coefficient: -0.200, 95% CI: [-0.389, -0.012], P = 0.04). Moreover, number of awakenings was associated with CV during the wake period of diastolic blood pressure (β-coefficient: -0.293, 95% CI: [-0.583, -0.003], P = 0.048, Table S1).

**Table 3.** Associations between simultaneous sleep variables and SBP outcomes.

|  | Total Sleep time/min | Sleep efficiency/% | Wake after sleep onset/min | Sleep onset latency/min | Number of Awaken |
| --- | --- | --- | --- | --- | --- |
| 24-hour Mean | 0.013 (-0.161, 0.186) | 0.392 (-1.248, 2.031) | 0.061 (-0.153, 0.275) | 0.398 (-1.330, 2.126) | 0.088 (-0.848, 1.024) |
| 24-hour SD | 0.000 (-0.054, 0.054) | -0.123 (-0.633, 0.386) | 0.000 (-0.068, 0.067) | 0.441 (-0.016, 0.899) | -0.191 (-0.453, 0.072) |
| 24-hour CV | -0.002 (-0.039, 0.036) | -0.121 (-0.469, 0.227) | -0.005 (-0.052, 0.041) | 0.323 (0.014, 0.632)* | -0.164 (-0.334, 0.006) |
| Mean in wake period | -0.001 (-0.176, 0.173) | 0.422 (-1.219, 2.064) | 0.066 (-0.148, 0.280) | 0.514 (-1.207, 2.235) | 0.070 (-0.869, 1.010) |
| SD in wake period | 0.014 (-0.050, 0.078) | -0.157 (-0.761, 0.447) | -0.008 (-0.088, 0.072) | 0.456 (-0.114, 1.027) | -0.236 (-0.545, 0.073) |
| CV in wake period | 0.010 (-0.032, 0.053) | -0.145 (-0.541, 0.252) | -0.012 (-0.065, 0.041) | 0.309 (-0.069, 0.686) | -0.200 (-0.389, -0.012)* |
| Mean in sleep period | 0.052 (-0.141, 0.244) | 0.233 (-1.626, 2.092) | 0.053 (-0.190, 0.295) | -0.011 (-1.976, 1.954) | 0.161 (-0.887, 1.209) |
| SD in sleep period | -0.014 (-0.050, 0.022) | -0.191 (-0.525, 0.144) | 0.025 (-0.019, 0.069) | 0.212 (-0.137, 0.561) | 0.038 (-0.162, 0.239) |
| CV in sleep period | -0.017 (-0.054, 0.020) | -0.183 (-0.534, 0.169) | 0.018 (-0.029, 0.066) | 0.199 (-0.169, 0.568) | 0.022 (-0.187, 0.231) |
| Weighted SD | 0.004 (-0.044, 0.052) | -0.153 (-0.601, 0.294) | 0.002 (-0.058, 0.062) | 0.371 (-0.045, 0.787) | -0.146 (-0.387, 0.094) |
| Weighted CV | 0.001 (-0.032, 0.034) | -0.144 (-0.442, 0.154) | -0.003 (-0.044, 0.038) | 0.272 (-0.003, 0.548) | -0.128 (-0.283, 0.027) |

The linear models were adjusted for age (continuous, years), sex (male, female), BMI, the duration of diabetes (continuous, years), antidiabetic medication use (oral medication only, oral medication and insulin injection), hypertension status and IPAQ MET (min/week). The results are shown as coefficient (95% CI), and statistical significance was indicated with asterisks.

### 3.3 Associations between multi-day sleep parameters and ABPM outcomes

The relationship between multi-day sleep parameters and ABPM outcomes are shown in **Table 4**. No significant associations were found between multi-day sleep parameters and systolic blood pressure variables, whereas multi-day sleep efficiency was associated with nighttime SD and CV of DBP (SD in sleep period of diastolic blood pressure: β-coefficient: -0.373, 95% CI: [-0.676, - 0.071], P = 0.02; CV in sleep period of diastolic blood pressure: β-coefficient: -0.588, 95% CI: [- 0.1.141, -0.036], P = 0.04, **Table S2**). In addition, the associations of hypertension types and dipping patterns with multi-day sleep-related variables were showed in **Figure S1**, with no significant differences observed.

**Table 4.**
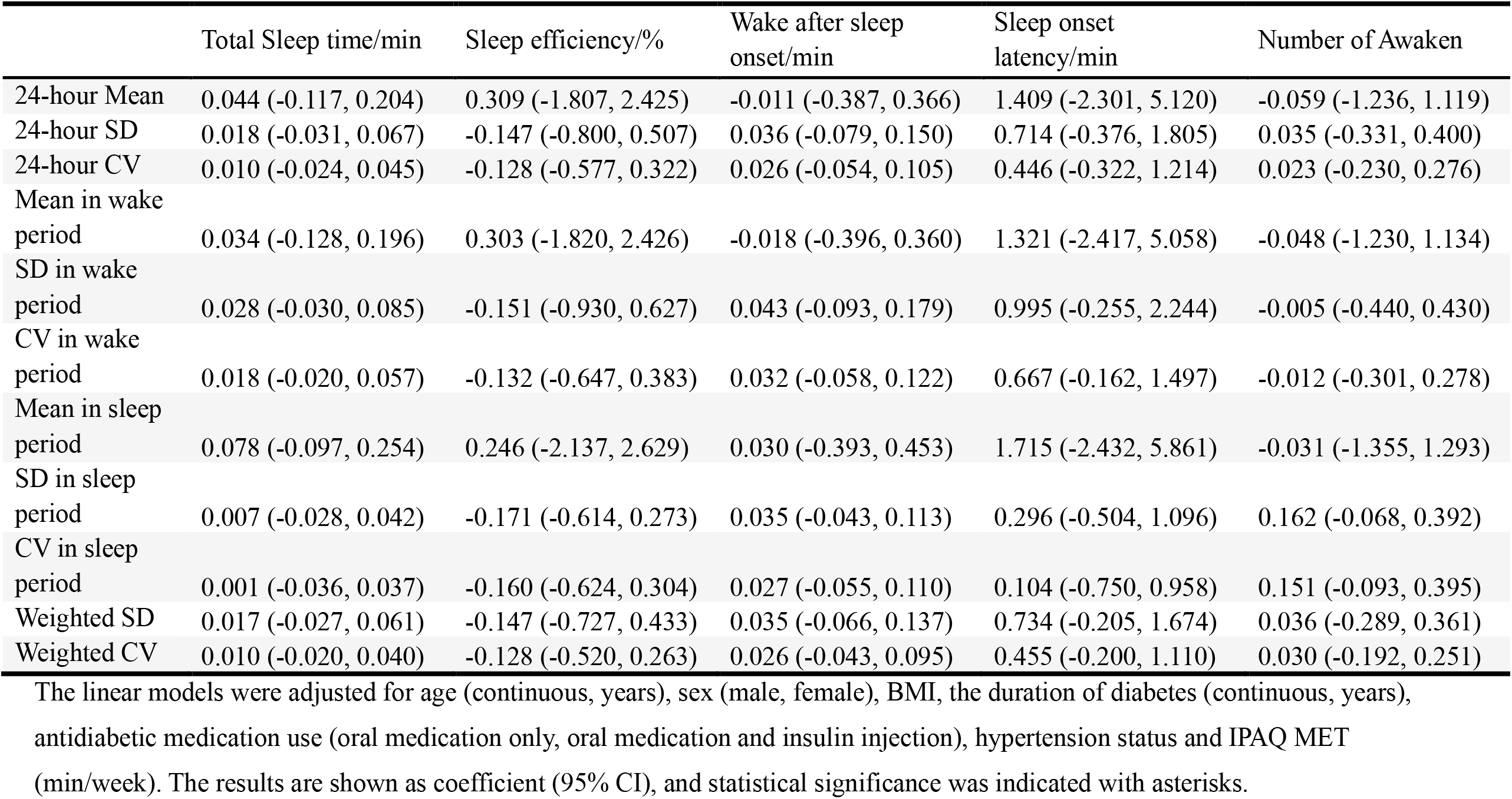
Associations between 14-days sleep variables and SBP outcomes.

## 4. Discussion

This study explored the associations between objective sleep parameters and 24-hour ABPM outcomes in individuals with T2D comorbid with hypertension. These findings suggest a notable association between blood pressure variability and key sleep characteristics, emphasizing the potential role of sleep in blood pressure regulation. The results demonstrated that specific sleep metrics were associated with blood pressure variability, particularly sleep onset latency and multi-day sleep efficiency. Sleep onset latency was correlated with higher systolic blood pressure CV over a 24-hour period, whereas reduced multi-day sleep efficiency was associated with greater diastolic blood pressure variability during the sleep period. These findings suggest that both acute and chronic disruptions in sleep quality are relevant to blood pressure regulation in T2D individuals.

Prolonged sleep onset latency is often a hallmark of sleep-onset insomnia, a condition characterized by difficulty initiating sleep despite adequate opportunity (22). This condition is particularly prevalent in individuals with T2D due to metabolic dysregulation, increased psychological distress, and pain-related comorbidities such as neuropathy (23, 24). The physiological consequences of extended sleep onset latency include increased sympathetic nervous system (SNS) activity, hypercortisolemia, and elevated arousal levels, all of which have been implicated in abnormal cardiovascular regulation (25). The elevated 24-hour SBP variability observed in our study may thus be partially mediated by sustained sympathetic activation and a reduced ability of the baroreflex to buffer rapid BP changes. In T2D patients, who often present with impaired autonomic regulation and diminished heart rate variability, these physiological disturbances may be further amplified, exacerbating cardiovascular strain (26). Furthermore, behavioral and psychological factors such as poor sleep hygiene, late-night eating, screen exposure, and underlying anxiety or depression may extend sleep onset latency, adding complexity to the relationship between sleep onset and BP control (27, 28). It was found that a 10-minute increase in sleep onset latency is correlated with 89% higher risk of hypertension (29), and a cross-sectional study found that longer sleep onset latency existed in hospitalized patients and was independently positively correlated with the risk of hypertension (30). A population-based study has shown that Swedish women with hypertension tend to exhibit longer sleep onset latency (31). These findings support the importance of early identification and management of sleep initiation difficulties as part of cardiovascular risk reduction strategies in patients with T2D.

Chronic sleep disturbances, particularly diminished multi-day average sleep efficiency, were also found to be associated with elevated DBP variability during the sleep period. Sleep efficiency, defined as the proportion of time spent asleep relative to the total time in bed, is a robust indicator of sleep continuity and quality. Reduced sleep efficiency is indicative of sleep fragmentation, characterized by frequent awakenings, increased WASO, and decreased restorative slow-wave sleep (32). From a physiological perspective, fragmented sleep interferes with the normal nocturnal decline in sympathetic activity and the corresponding rise in parasympathetic tone, which are essential for cardiovascular recovery and BP stabilization during sleep (33). Consequently, disrupted autonomic balance during sleep results in fluctuating vascular resistance and hemodynamic instability, manifesting as greater DBP variability. Diastolic pressure, particularly during sleep, reflects peripheral vascular resistance and is closely regulated by endothelial function and smooth muscle tone. In individuals with T2D, who often exhibit endothelial dysfunction, insulin resistance, and low-grade inflammation, these regulatory systems are already compromised. The compounding effect of poor sleep further exacerbates these mechanisms, leading to increased nocturnal BP variability. Moreover, evidence suggests that poor sleep efficiency may interfere with melatonin secretion—a hormone with vasodilatory and chrono-biotic properties that supports nocturnal BP reduction (34). The attenuation of melatonin release in individuals with disrupted sleep may lead to blunted dipping and increased vascular tone, contributing to adverse cardiovascular remodeling over time. Our findings emphasize the role of chronic sleep quality, not just duration, in shaping blood pressure dynamics and highlight the potential for behavioral sleep interventions to stabilize nocturnal BP fluctuations in hypertensive T2D patients.

The observed association between sleep disturbances and blood pressure variability carries significant metabolic and cardiovascular implications for individuals with T2D (35). T2D is commonly accompanied by autonomic dysfunction, endothelial impairment, and heightened inflammatory responses—all of which contribute to the disruption of circadian blood pressure regulation (36, 37, 38). Abnormal nocturnal blood pressure patterns, including reduced or reversed dipping, are not only linked to increased cardiovascular risk but also strongly associated with greater insulin resistance, potentially mediated by heightened sympathetic activity, disrupted cortisol rhythms, and increased hepatic glucose output (39, 40). These pathophysiological processes may establish a vicious cycle of “sleep disruption–blood pressure dysregulation–metabolic imbalance”. Therefore, in the clinical management of T2D, the integration of objective sleep assessments with ambulatory blood pressure profiles may provide a more comprehensive framework for cardiovascular risk stratification and offer new avenues for preventing metabolic complications.

This study has several notable strengths. First, it is a study to integrate actigraphy-based sleep assessment with 24-hour ambulatory blood pressure monitoring in patients with T2D comorbid with hypertension. This dual-method approach allowed for objective and complementary measurements of both sleep and blood pressure dynamics, reducing the reliance on self-reported data and increasing the accuracy of results. Second, the study adopted a multi-day sleep monitoring protocol (14 consecutive days), which enhanced the ecological validity of the sleep measures and captured sleep patterns rather than relying on single-night assessments that may be affected by night-to-night variability. Importantly, this study examined both concurrent sleep (matched to the ABPM night) and aggregated multi-day sleep parameters, which provided complementary insights into acute versus chronic sleep–blood pressure associations. Finally, it focused on individuals with T2D and hypertension who are at particularly high risk for cardiovascular complications, thereby underscoring the translational value of the findings for patient care and risk management.

Despite these strengths, several limitations should also be acknowledged and should be addressed in future research. First, the observational nature and relatively small sample size limit the statistical power and generalizability of the results. Although strict inclusion criteria and objective measurements were employed, the limited representativeness of the sample underscores the need for validation in larger and more diverse populations. Second, the cross-sectional design precludes any conclusions regarding the causal relationship between sleep parameters and blood pressure indices. For instance, elevated nocturnal blood pressure may result from disrupted sleep, but it could also contribute to poor sleep quality, suggesting the need for longitudinal and intervention studies to clarify the directionality of these associations. Third, while adjustments were made for several important confounders (e.g., age, sex, BMI, diabetes duration, physical activity), unmeasured factors such as depressive symptoms, medication adherence, and dietary habits may still influence both sleep and blood pressure outcomes. Lastly, the study did not employ polysomnography (PSG) thereby precluding analysis of specific sleep disorders such as obstructive sleep apnea, which may significantly contribute to nocturnal hypertension. Despite these limitations, the present study highlights critical associations between sleep characteristics and ambulatory blood pressure profiles in T2D patients, underscoring the potential of sleep-targeted interventions to improve circadian blood pressure regulation and mitigate cardiovascular risk. Future studies should employ longitudinal designs with larger cohorts, incorporate comprehensive physiological and behavioral assessments, and evaluate the efficacy of tailored interventions to inform personalized care strategies.

## Data Availability

Dr. Xiao Tan is the guarantor of this work and, as such, had full access to all the data in the study and takes responsibility for the integrity of the data and the accuracy of the data analysis.

## Ethics approval and consent to participate

Ethics approval was obtained from both Ethics Committees at Nanjing Sport Institute (RT-2023-17) and at Community Health Service of Qixia District, Nanjing Municipality (2023-QX-040). The trial was registered at ClinicalTrials.gov (NCT06145542). Written informed consent were obtained from all participants prior to baseline measurements.

## Acknowledgments

The authors thank the participants and researchers who contributed or collected data. Dr. Xiao Tan is the guarantor of this work and, as such, had full access to all the data in the study and takes responsibility for the integrity of the data and the accuracy of the data analysis.

## Funding

This research was funded by Jiangsu provincial key research and development program (Y.Z., BE2022828), National Natural Science Foundation of China (X.T., 82570128), Development Project of Zhejiang Province (X.T., 2025C01119), Rut and Arvid Wolff Memorial Foundation (X.T., 2023-02467), and Jiangsu provincial key research and development program (Y.Z., BE2022828). The funders had no role in the conduct of the study; collection, management, analysis, or interpretation of the data; preparation, review, or approval of the manuscript; or decision to submit the manuscript for publication.

## Declaration of interests

There are no conflicts of interest to disclose.

## Contribution Statement

Yan Zhao: Conceptualization, Methodology, Supervision, Project administration, Funding acquisition;

Yuzhe Zha: Investigation

Yuchan Zheng: Investigation

Lijun Wei: Investigation

Baoyi Chen: InvestigationÍ

Zefan Zhang: Investigation, Formal analysis, Writing – Original Draft, Writing – Review & Editing, Visualization;

Xiao Tan: Conceptualization, Formal analysis, Investigation, Supervision, Funding acquisition, Writing – Original Draft, Writing – Review & Editing, Project administration.

## Notes

### Competing Interest Statement

The authors have declared no competing interest.

### Clinical Trial

NCT06145542

### Author Declarations

Ethics approval was obtained from both Ethics Committees at Nanjing Sport Institute (RT-2023-17) and at the Community Health Service of Qixia District, Nanjing Municipality (2023-QX-040).

